# Investigation of the Relationship between Delta Inflammatory Markers and Prognosis in Head and Neck Squamous Cell Carcinoma

**DOI:** 10.1101/2021.04.29.21256130

**Authors:** Julian Khaymovich, Andrew Ko, Amanda Wong, Daniel Zhu, Christian Gigante, Srilakshmi Garikapati, Ginnie Jeng, Sarah Van Der Elst, Charles Rong, Tristan Tham

## Abstract

**Purpose:** Inflammatory markers, such as Lymphocyte-to-Monocyte Ratio (LMR), Neutrophil-to-Lymphocyte Ratio (NLR), and Platelet-to-Lymphocyte Ratio (PLR), have been shown to hold significant prognostic value in the context of head and neck cancer (HNC). Recently, delta inflammatory markers, the difference between pre and post- treatment inflammatory marker ratios, have been suggested as potentially significant values in predicting cancer prognosis. Our objective was to evaluate the prognostic utility of delta LMR, NLR, and PLR in head and neck squamous cell carcinoma (HNSCC).

**Methods:** Retrospective cohort study in a tertiary academic hospital setting. Patients diagnosed with HNSCC in the oral cavity, larynx, and oropharynx treated with curative intent treatment were included. The variables collected were age, sex, BMI, alcohol/tobacco exposure, performance scores, ACE-27, tumor characteristics, adjuvant treatment, ECOG score, and lab values. Overall Survival (OS) and Event-Free Survival (EFS) were chosen as endpoints. OS was defined as time from date of treatment to date of last follow-up or death from any cause, and EFS was defined as the start of treatment to any progression, recurrence, or death from any cause. Univariate and multivariate analyses were performed on the primary endpoints.

**Results:** A total of 89 patients were included from 2010 to 2017. In multivariate analysis, EFS was found to be significantly associated with an N stage of 3 (p=0.0005) and delta LMR > -1.48 (p=0.0241). No significant relationships were uncovered with OS in multivariate analysis.

**Conclusion:** A higher delta LMR value (>-1.48) was associated with poorer EFS, but was not associated with OS.

## Introduction

Squamous Cell Carcinoma of the Head and Neck (HNSCC) is the largest and most commonly occurring subset of Head and Neck Cancer (HNC), appearing in the squamous epithelium of the oral cavity, larynx, and oropharynx [1, 2]. Treatment indication is based on diagnostic workup, which ultimately depends on several factors that are widely accepted as indicators of prognosis in patients with HNSCC. The classic prognostic factors include tumor differentiation, site, size, nodal involvement, metastasis, tumor grading, in addition to age and various comorbidities [3–6]. Smoking and alcohol usage status have also classically been useful in indicating tumor prognosis. More recently, HPV status has become an important factor to consider in HNSCC prognosis, playing a significant role in tumor growth via subsequent degradation of the p53 tumor suppressor gene [7, 8]. Lately, there has also been increased interest in the use of inflammatory biomarkers as prognostic indicators for HNC.

The neutrophil-to-lymphocyte ratio (NLR), lymphocyte-to-monocyte ratio (LMR), and platelet-to-lymphocyte ratio (PLR) have all been shown to be potentially reliable values in the prediction of HNC outcomes [9–15]. Markers of systemic inflammation have been shown to be associated with tumor stage and progression, likely due to inflammatory reactions throughout tumor development [16]. For example, a recent observational study showed that elevated NLR and PLR in HNC patients treated with chemoradiotherapy or surgery significantly correlated with poor overall survival [17]. In addition, the monitoring of NLR values in patients with recurrent or metastatic HNSCC undergoing chemotherapy with nivolumab can be a significant predictor of treatment failure [18]. In laryngeal cancer patients treated with induction chemotherapy, a high pre-treatment NLR was shown to be associated with poorer survival, independent of the response to chemotherapy [19].

These findings have shown the predictive significance of the monitoring of inflammatory values throughout the course of treatment, pointing to the use of the delta (Δ) ratio (pre-treatment subtracted from post-treatment value) as a potential prognostic indicator. In the past, the delta ratio has been used in other cancers such as colon cancer and has shown delta NLR to be an independent prognostic indicator of overall survival [20]. In addition, delta NLR has been shown to be an independent predictor of overall survival in patients with advanced non-small cell lung cancer [21]. Delta NLR has also proven itself to be a strong predictor in breast cancer. Delta NLR, rather than solely pre-treatment or post-treatment values, was associated with pathological complete response to neoadjuvant chemotherapy in breast cancer patients [22]. In glioblastoma patients undergoing laser interstitial thermal therapy, larger delta NLR were associated with greater overall survival [23]. However, there are fewer studies that focus on the use of such values in patients with HNSCC. Therefore, the aim of our study was to look at the delta NLR, delta PLR, and delta LMR in patients that have undergone treatment for HNSCC. These values are easily obtainable and affordable, further adding to their potential utility as prognostic factors.

## Materials and Methods

### Study Design

This retrospective cohort study included patients with HNC treated at the Northwell Health System from 2010 to 2017. This study was approved by the Institutional Review Board of the Northwell Health System (IRB#19-0336). The inclusion criteria for the study was (a) histologically confirmed Squamous Cell Carcinoma of the Head and Neck (HNSCC), (b) documentation of curative intent treatment with surgery as the primary modality of treatment, with or without adjuvant therapy, (c) availability of complete clinical data and disease records (pre-operative and post-operative data), and (d) a location in the oral cavity, larynx (including supra/subglottis and all sites associated with larynx), and oropharynx.

The exclusion criteria was (a) an incomplete documentation of curative treatment, (b) lack of treatment or palliative treatment, (c) biopsies or non-curative intent procedures, (d) metastatic disease at time of presentation, (e) presence of tumor in nasopharynx, nasal cavity, paranasal sinus, (f) an unknown primary tumor (T0), (g) presence of carcinoma-in-situ (cis), and (h) a histology other than HNSCC. Patients were screened using pre-defined ICD-9/10 codes. All patients were treated according to plans that were consistent with contemporary treatment paradigms [24].

### Variable Selection

Established prognostic factors for HNC were included in the data collection process: sex, age ethnicity, BMI, tumor site, tumor differentiation, tumor stage according to the 8th Edition of the American Joint Committee on Cancer (AJCC), Eastern Cooperative Oncology Group (ECOG) score, smoking history, alcohol history, and comorbidities based upon Adult Comorbidity Evaluation Score-27 (ACE-27) [25]. Both pre-treatment, defined as anywhere within a month before treatment, and post-treatment, defined as within one to three months after treatment, were obtained. The pre-treatment and post-treatment NLR were defined as the neutrophil count divided by the lymphocyte count. The pre-treatment and post-treatment LMR count was defined as the lymphocyte count divided by the monocyte count. The pre-treatment and post-treatment PLR were defined as the platelet count divided by the lymphocyte count. The delta (Δ) ratio was defined as the pre-treatment ratio subtracted from the post-treatment ratio. For example, the Δ LMR was defined as pre-treatment lymphocyte count divided by pre-treatment monocyte count subtracted from post-treatment lymphocyte count divided by the post-treatment monocyte count. The electronic healthcare records of patients were reviewed and stored in a REDCap database [26]. Clinical information was retrieved from the patient’s scanned notes and relevant clinical, pathological, or laboratory reports.

### Endpoint

Overall Survival (OS) and Event-Free Survival (EFS) were chosen as endpoints. OS was defined as time from date of treatment to date of last follow-up or death from any cause. Patients who did not experience the event of interest (death) were censored. EFS was defined as the start of treatment to the last follow-up or ‘event’. An ‘event’ was defined as any progression, local or distant recurrence, or death from any cause. Patients who did not experience any ‘event’ were censored.

### Statistical Analysis

To find the optimal cutoff points for the continuous variables NLR, LMR, PLR, and their delta values, we used the construction of a receiver operator characteristic curve (ROC plot) which is a plot of the sensitivity against 1-specificity, of the detection of the endpoint (EFS). After the ROC curve is constructed, the discrimination threshold of the continuous variable is identified using the Youden index, which is the level at which the continuous variable maximizes both sensitivity and specificity. Using this method, separate cutoff estimates were obtained for all continuous variables of interest (NLR, LMR, PLR, and their delta values) for EFS. For example, an ROC curve for delta LMR can be seen in Figure 1. The Kaplan–Meier curve (KM) was used to estimate survival endpoints, and the logrank test was used to find survival differences between KM curves. Additionally, we used the Cox proportional hazards model (CPH) to determine the hazard ratio (HR) of the variables listed above. Univariate and multivariate analyses were performed for EFS. If a variable was significant on univariable analysis, it was selected for the multivariable model. In generating the multivariate CPH model, backwards variable selection was used. The threshold for removing a variable from the backwards selection multivariate model was set to p=0.1. Univariate p values are reported, and multivariate p values>0.1 are reported as ‘ns’ (non-significant). HRs with 95% confidence intervals (95% CI) are presented, with two-sided p values. The alpha level was set to 0.05, and p values less than 0.05 were considered statistically significant. All statistical analyses were performed using MedCalc for Windows, version 15.0 (MedCalc Software, Ostend, Belgium).

**Figure 1.**
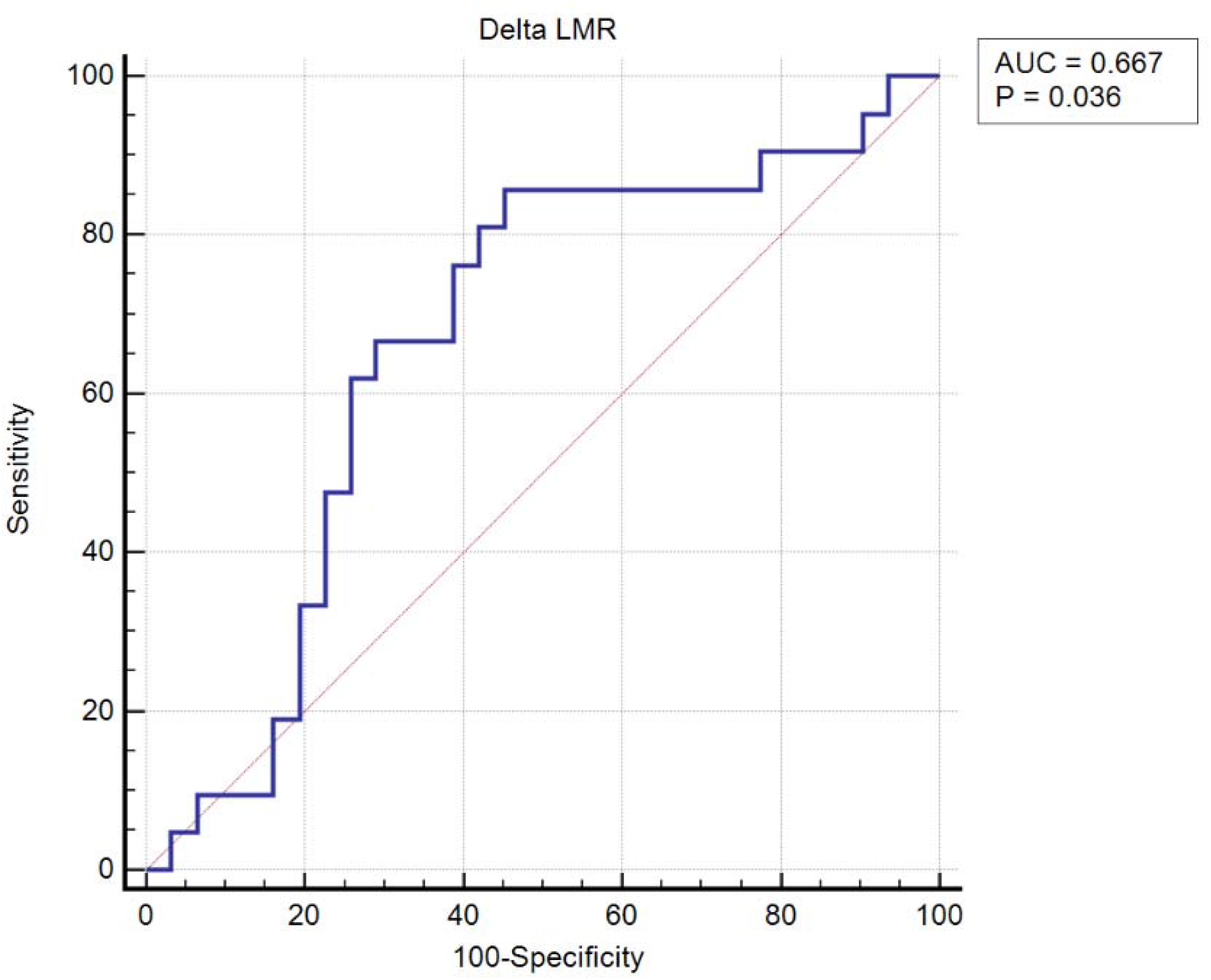
To find the optimal cutoff points for the continuous variables NLR, LMR, PLR, and their delta values, we used the construction of a receiver operator characteristic curve (ROC plot) which is a plot of the sensitivity against 1-specificity, of the detection of the endpoint (EFS). This figure shows the ROC curve for delta LMR.

## Results

A total of 89 patients were enrolled in this study, including 58 males and 31 females. The primary tumor sites of the squamous cell carcinoma were: oropharynx (26), oral cavity (45), hypopharynx (2), and larynx (16). Among the tumor cases, 24 were poorly differentiated, 42 were moderately differentiated, 11 were well differentiated; the remaining cases were not reported. Treatment with radiotherapy was used in 66 of the patients, chemotherapy was used in 49 patients, and surgery was used in 59 patients. The clinical characteristics of the cohort can be found in Table 1.

**Table 1.** Clinical features and characteristics of study population

| Variables | Total (n=89) |
| --- | --- |
| Sex |  |
| Male | 58 |
| Female | 31 |
| BMI (kg/m<sup>2</sup>) | 27.2 ± 5.6 |
| Alcohol history |  |
| None | 34 |
| Active | 42 |
| Active/Abuse | 2 |
| Quit | 9 |
| Unknown | 2 |
| Smoking history |  |
| None | 38 |
| Active | 10 |
| Quit | 40 |
| Unknown | 1 |
| ECOG score |  |
| 0 | 44 |
| 1 | 35 |
| 2 | 2 |
| 3 | 0 |
| Karnofsky score (%) | 86.3 ± 6.1 |
| ACE-27 score |  |
| 0 | 16 |
| 1 | 50 |
| 2 | 14 |
| 3 | 9 |
| Tumor differentiation |  |
| Poor | 24 |
| Moderate | 42 |
| Well | 11 |
| Not reported | 6 |
| Tumor site |  |
| Oropharynx | 26 |
| Oral Cavity | 45 |
| Larynx | 16 |
| Hypopharynx | 2 |
| T stage |  |
| T1, T2 | 50 |
| T3, T4 | 39 |
| N stage |  |
| N0 | 36 |
| N1+ | 53 |
| HPV Status |  |
| Positive | 25 |
| Negative/Unknown | 64 |
| Treatment modality |  |
| RT | 66 |
| CT | 49 |
| Surgery | 59 |

BMI: body mass index, ECOG: eastern cognitive group, ACE-27: adult comorbidity evaluation score 27, HPV: human papillomavirus, RT: radiation therapy, CT: chemotherapy

Univariate analysis of OS showed significant correlation with BMI (p=0.0082, HR= 0.84, 95% CI, 0.74 to 0.96), a primary tumor location of the hypopharynx (p=0.0118, HR= 37.85, 95% CI, 2.24 to 640.94), advanced T stage (T4) (p=0.024, HR= 6.71, 95% CI, 1.29 to 35.01), unknown history of alcohol use (p=0.0005, HR= 22.79, 95% CI, 3.90 to 133.15), and ECOG score (p=0.0334, HR= 3.61, 95% CI, 1.11 to 11.75). Inflammatory markers were not statistically significant towards differences in OS, so multivariate analysis was not performed for the significant variables.

Univariate analysis of EFS revealed significant correlation with BMI (p=0.0254, HR=0.92, 95% CI, 0.85 to 0.99), an N stage of 3 (p=0.0003, HR= 7.29, 95% CI, 2.48 to 21.40), pre-treatment LMR (p=0.0169, HR= 3.11, 95% CI, 1.23 to 7.90), delta LMR > -1.48 (p=0.0125, HR=4.75, 95% CI, 1.40 to 16.15), delta NLR < 0.18 (p=0.0119, HR= 3.05, 95% CI, 1.28 to 7.28), an ECOG score of 1 (p= 0.0260, HR= 2.32, 95% CI, 1.11 to 4.86), and unknown history of alcohol use (p=0.0002, HR= 23.85, 95% CI, 4.48 to 126.82). Univariate and multivariate results are illustrated in Table 2. In order to control for covariates, multivariate analysis was performed. The following variables were found to be significantly associated with EFS in the multivariate model: an N stage of 3 (p=0.0005, HR= 7.36, 95% CI, 2.39 to 22.70) and delta LMR > -1.48 (p=0.0241, HR= 4.17, 95% CI, 1.21 to 14.41). A Kaplan-Meier curve was constructed and a log rank test showed a significant difference between delta LMR values above and below -1.48 (p=0.0058, Figure 2).

**Table 2.**
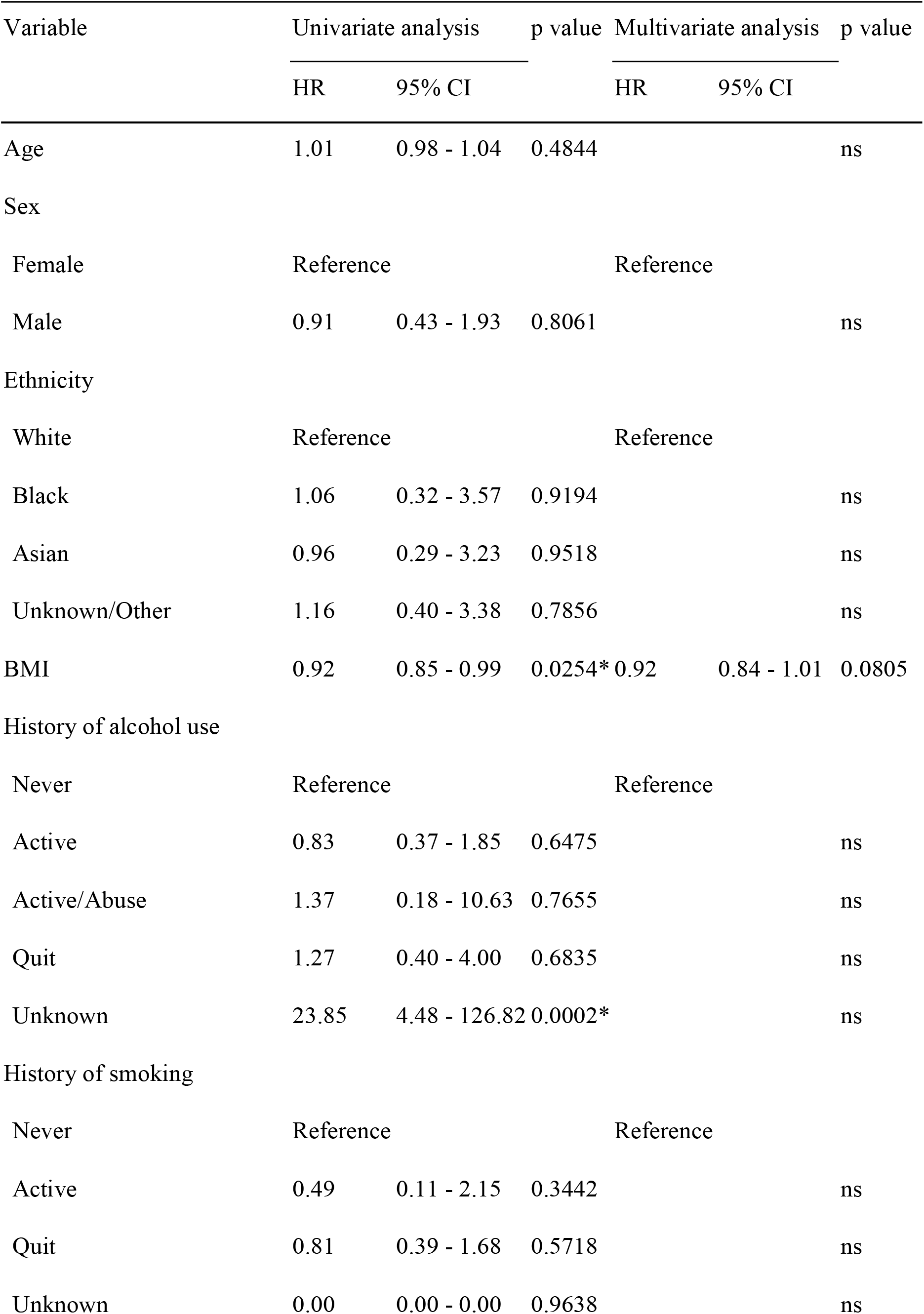
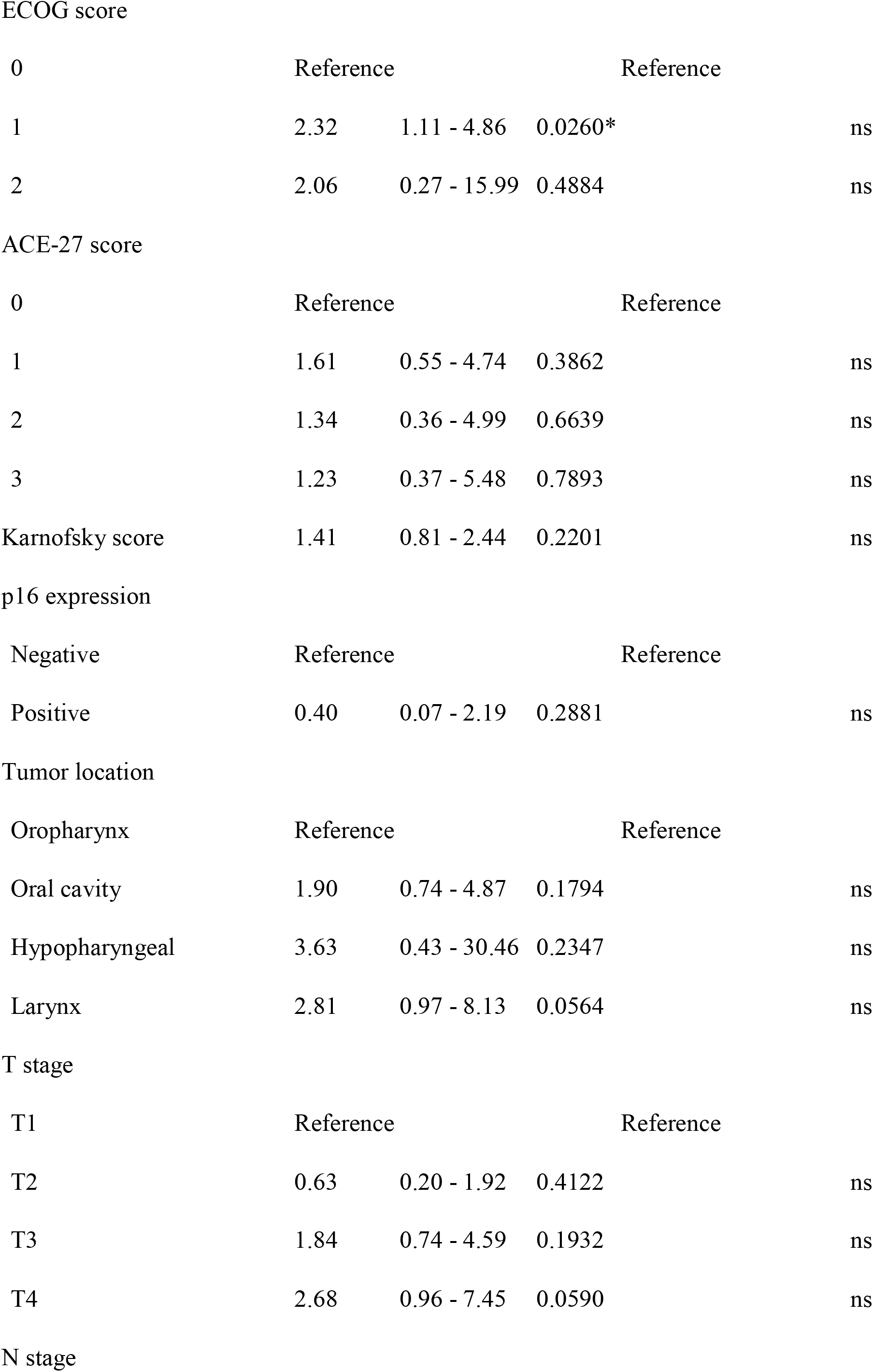
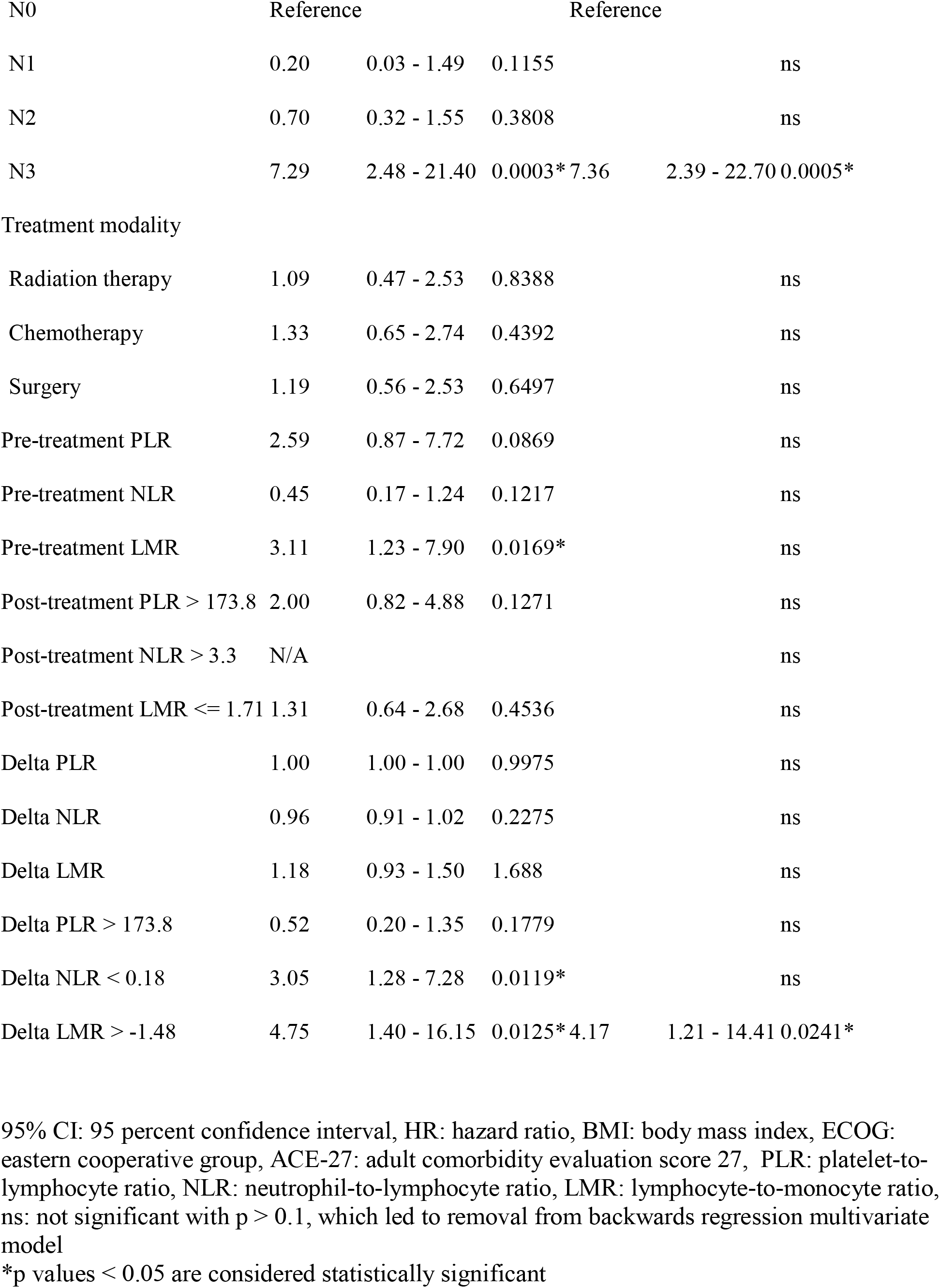
Univariate and multivariate analysis of event-free survival (EFS)

**Figure 2.**
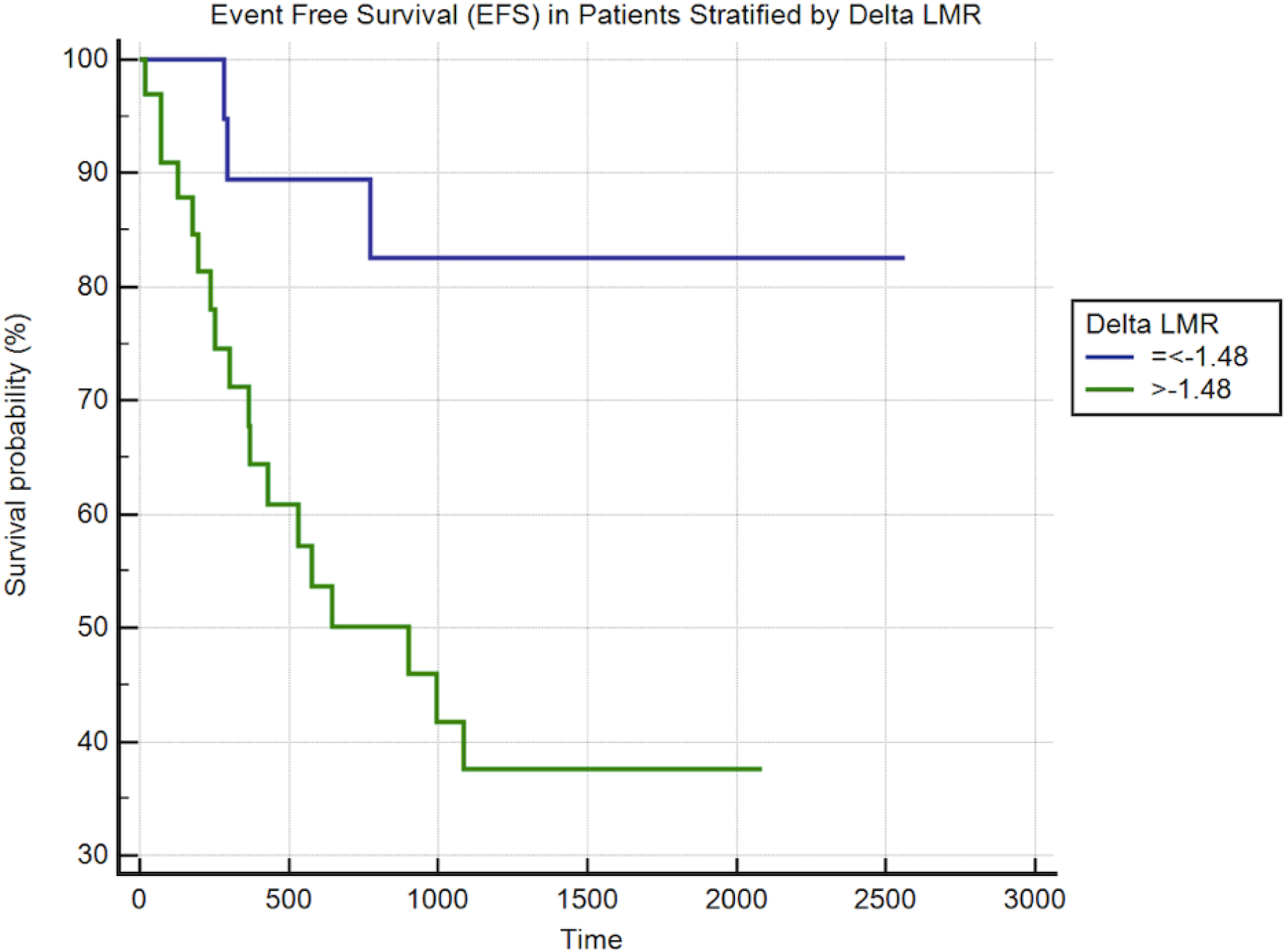
Event Free Survival (EFS) in Patients Stratified by Δ LMR. Based on a log rank test, the difference in survival between groups was significant (p = 0.0058). Patients with lower delta LMR (blue) had better EFS outcomes than those with higher delta LMR (green).

## Discussion

It has long been known that inflammation plays an important role in tumor progression through various pathways and mechanisms [27]. The tumor micro-environment of HNSSC involves a diverse array of cells, including neutrophils, monocytes, and platelets, among many others. Neutrophils appear early in the immune response, acting as a primary defense. T-lymphocytes act as a key component of antitumor activity; however, as oncogenesis continues, this lymphocytic response is suppressed and apoptosis increases, leading to rapid turnover [28]. In the meantime, tumor associated macrophages (TAMs) are a major part of the infiltrating leukocyte response. The macrophages arrive at the site of the neoplasm in large amounts and play a significant role in tumor growth and lymph node metastasis, secreting factors that encourage angiogenesis and tumor aggressiveness, and evasion of immune surveillance [29]. The macrophages produce IL-8, for example, which further signals an inflow of inflammatory cells. In addition, cancer is associated with a prothrombotic state, due to procoagulant factors being expressed by tumor cells. Platelet count and turnover resultantly increases [30]. The tumor micro-environment includes numerous interactions between various factors and cells, leading to a complex response that is still not fully understood. The development of a neoplasm begins with an acute inflammatory response, and proceeds to a state of chronic inflammation as the oncogenesis is unable to be halted. Thus, the cells involved in the inflammatory process potentially hold significant value in the determination of the prognosis of a tumor.

In the present study, a noteworthy finding was that delta LMR was significantly associated with EFS, but not OS. Patients with a lower delta LMR (delta LMR< -1.48) maintained a better prognosis than those with a higher delta LMR (delta LMR >-1.48) (p=0.0241, HR= 4.17, 95% CI, 1.21 to 14.41), as can be seen in Figure 2. This finding supports current available literature. In a large study by Lin et al., longitudinal analysis of inflammatory markers throughout radiotherapy treatment in 1,431 patients with HNC showed a significant association between delta LMR and overall survival (OS) and freedom from metastasis (FFM), with improved survival amongst patients with lower delta LMR [5]. Unlike the study by Lin and colleagues, in which only radiotherapy treatment was evaluated, our study evaluated delta prognostic factors with several different treatment modalities, including radiotherapy. Although we did not identify a significant relationship between OS and delta LMR in our analysis, this study provides supporting evidence of a relationship between survival and delta LMR. This relationship is potentially due to lymphocyte and macrophage roles within the tumor microenvironment. Lymphocyte presence has been associated with favorable outcomes, while macrophages have been associated with tumor progression [31, 32]. Therefore, a ratio presents the ability to evaluate their relationship. The use of serial LMR can also explain treatment-induced changes in immune function by capturing changes in immune status due to radiotherapy, for example [33]. The high predictive value of delta LMR over static inflammatory markers suggests its potential use as a useful prognostic factor.

According to univariate analysis, we found that delta NLR less than 0.18 had a significantly worse prognosis of EFS (p=0.0119, HR= 3.05, 95% CI, 1.28 to 7.28), in addition to pre-LMR also maintaining a significant relationship with EFS (p=0.0169, HR= 3.11, 95% CI, 1.23 to 7.90). Although these results did not prove to be significant in multivariate analysis, they are supported by current literature studying the use of delta inflammatory biomarkers in non-small-cell lung cancer (NSCLC) [34]. Delta NLR and pre-LMR were significantly associated with OS and progression-free survival (PFS), in both univariate and multivariate analysis in patients with NSCLC undergoing concurrent chemoradiotherapy. While delta inflammatory biomarkers did not prove to be significant in OS in our findings, the slight differences in results can be explained by several reasons. First, our study incorporated several treatment modalities, including chemotherapy, radiotherapy, or surgery, rather than a single modality. Second of all, our study specifically evaluated HNSCC, rather than NSCLC, and the varying immune response to different tumors may explain the subsequent differences in findings.

In addition to inflammatory markers, other clinical characteristics were found to be significant predictors of HNSCC prognosis. An N stage of 3, according to the TNM classification model, was found to be highly correlated with poor EFS upon multivariate analysis. Univariate analysis showed several independent relationships among BMI, ECOG score, unknown alcohol use history, advanced T stage (T4), tumor location within the hypopharynx, and OS. BMI, advanced N stage (N3), ECOG score, and unknown alcohol history were independent predictors of EFS. These findings are supported by current literature. Advanced N and T stages have both been shown to significantly worsen prognosis, due to the nature of tumor invasion and metastasis in late stages [35, 36]. Higher BMI, for example, was found to have a survival advantage in our study, in terms of OS (p=0.0082, HR= 0.84) and EFS (p=0.0254, HR=0.92). These patients are thought to have an increased capacity in tolerating cancer treatments, resulting in improved prognosis [37, 38]. Alcohol use has also been found to negatively affect HNC development and prognosis, supporting our findings as well [39].

Despite developing novel findings in our study, we would also like to acknowledge its shortcomings. First, this was a single center study which was conducted as a retrospective cohort study. As a result, it may have been subject to retrospective bias. In addition, although data collection points were limited, they were not standardized, potentially altering results (i.e., post-treatment data was collected anytime from one to three months post-treatment). Furthermore, as a result of the specificity of the inclusion and exclusion criteria, our sample size was relatively small. Because our sample size was small, we were also unable to stratify according to HNC subsite, however we were able to control for subsite in the multivariate analysis. In addition, previous studies have found that the effect of inflammatory markers is consistent across separate HNC sites [40].

In conclusion, using a multivariate prognostic model, delta LMR was found to be a significant prognostic factor of EFS. In order to further validate the results this study presents, future high-quality prospective studies, with larger sample sizes, must be performed. The development of these novel prognostic factors might improve prognostic stratification in this era of individualized treatment planning.

## Data Availability

N/A

## Acknowledgments

We would like to thank Yuna Choi, Josue Minaya, Arif Mahmud, and Zachary Lowy for assistance in the data collection process.

## Ethics Approval

N/A

## Consent to participate

N/A

## Consent for publication

N/A

## Availability of data and material

N/A

## Code availability

N/A

